# Variance polygenic scores (vPGS) as a tool for studying gene-environment interactions associated with refractive error

**DOI:** 10.64898/2026.05.06.26352553

**Authors:** Xi He, Louise Terry, Jeremy A. Guggenheim, The MyoTreat Network, UK Biobank Eye and Vision Consortium

## Abstract

**Purpose:** Conventional polygenic scores predict an individual’s phenotype based on their genetics. By contrast, variance polygenic scores (vPGS) quantify genetic predisposition to phenotypic variance. We tested the hypothesis that a vPGS for refractive error can identify individuals with increased susceptibility to environmental risk factors for myopia.

**Methods:** Six vPGS construction strategies were evaluated in UK Biobank participants: three variance heterogeneity genome-wide association studies (vGWAS) methods and two reweighting schemes. vPGS performance was assessed using two metrics: (i) ‘Diff’ – difference in phenotypic variance in vPGS decile one vs. ten; (ii) Spearman correlation of phenotypic variance vs. vPGS decile. The optimal vPGS was used to test for vPGS × time spent reading or vPGS × time spent outdoors interactions in children aged 15 years (ALSPAC cohort; n=3471).

**Results:** Of the vGWAS methods, conditional quantile regression outperformed SCAMPI and Levene’s Test. Of the re-weighting schemes, LDpred2 outperformed pruning and thresholding (P+T). In an independent sample of UK Biobank participants (n=19470), the top-performing vPGS successfully stratified individuals into groups with increasing variance in refractive error, even after adjusting for a conventional PGS (Diff: 2.55, 95% CI: 1.64–3.47; Spearman correlation: 0.87, 95% CI: 0.43–0.93). However, in ALSPAC participants, there was minimal support for vPGS interactions with time reading (P=0.80) or time outdoors (P=0.89).

**Conclusion:** A novel vPGS successfully stratified individuals into groups with relatively high or low genetic susceptibility to refractive error variance. However, the vPGS could not identify individuals at enhanced risk from lifestyle risk factors for myopia.

## Introduction

Both genetic and lifestyle risk factors confer susceptibility to refractive errors such as myopia and hyperiopia.^1^ Myopia is associated with myopic maculopathy, retinal detachment and glaucoma, while hyperopia is associated with strabismus and amblyopia.^2,3^ The prevalence of myopia is rising globally.^4^ Genome-wide association studies (GWAS) have been a powerful tool for investigating the genetic architecture of refractive error; to date, more than 450 common variants associated with the risk of myopia and hyperopia have been identified in GWAS analyses of more than 500,000 individuals.^5^ Excessive near work and insufficient time outdoors have been intensively studied as potential environmental risk factors for myopia.^6^ In addition to the risk conferred independently by genetic and environmental risk factors, existing evidence also supports a contribution from gene-environment interactions.^7^

Several genes have been implicated in gene-environment interactions associated with myopia development, including genes such as *LAMA2*, *GJD2* and *ZMAT4* that have been replicated in independent samples.^8–11^ Early gene-environment interaction studies of myopia tended to focus on candidate genes identified in GWAS analyses or based on biological function, since directly testing millions of genetic variants across the genome for evidence of a gene-environment interaction incurs a heavy multiple testing burden. More recently, variance heterogeneity genome-wide association studies (vGWAS) have been used successfully as an indirect route for finding gene-environment interaction loci. The variance quantitative trait loci (vQTL) identified in a vGWAS are genetic variants associated with a difference in trait variance across genotype categories, which is a characteristic feature of loci involved in gene-environment or gene-gene interactions. Levene’s test (LT) and closely-related methods such as the Brown-Forsythe test were originally implemented in vGWAS analyses.^12–15^ Subsequently, conditional quantile regression (CQR), which can identify a difference in genetic effect size across phenotype quantiles, has also shown promising results; CQR provides greater statistical power compared to LT in some circumstances.^16,17^ More recently still, a vGWAS method called SCAMPI (Scalable Cauchy Aggregate test using Multiple Phenotypes to test Interactions) has been reported.^18^ SCAMPI implements a variance-based algorithm similar to LT but considers variance heterogeneity across multiple correlated phenotypes to boost statistical power.^18^

Building on the concept of a conventional polygenic score (PGS), which is a method for predicting the phenotype of an individual based on their genetics, a small number of studies have instead explored the concept of a variance polygenic score (vPGS) for predicting an individual’s genetic susceptibility to phenotypic variance.^19–22^ Persons with a relatively high vPGS are expected to have greater “phenotype plasticity”. In a recent study^23^, we compared three vGWAS methods for detecting variance heterogeneity associated with refractive error: LT, CQR and SCAMPI. In the current work, we utilized the summary statistics from the three vGWAS analyses to construct an optimal vPGS and then tested the hypothesis that individuals with a relatively high vPGS were more susceptible to the effects of lifestyle risk factors for myopia.

## Methods

### Sample selection: UK Biobank participants

UK Biobank (UKB) recruited more than 500,00 participants, aged 40-70, from England, Scotland, Wales and Northern Ireland. Ethical approval for the study was obtained from the NHS Research Ethics Committee (Ref 11/NW/0382) and all participants provided written informed consent. At a baseline clinical assessment visit, approximately 25% UKB participants underwent non-cycloplegic autorefraction (Tomey RC 5000 autorefractor, Tomey Corp., Nagoya, Japan).

SER was calculated by the formula: SER = autorefraction sphere power + 0.5 × autorefraction cylinder power. SER was averaged between the right and left eyes to provide a phenotype value for each individual. If data for both eyes were not available, then the eye with the single record was used instead. All UKB participants were asked to self-report their age of onset of starting to wear spectacles or contact lenses (AOSW) and the age of completed full-time education (EduAge), except for those with a higher degree. Therefore, individuals with a university or college degree were categorized as completing 21 years of education. To account for low participant counts beyond EduAge values above 21 years and below 15 years, those who reported completing their full-time education before 15 years-old were recorded as having completed education at age 15 years, while those who reported completing their education beyond age 21 years were recorded as completing their education at age 21 years. As described previously,^24^ DNA samples from UK Biobank participants were genotyped using one of two single nucleotide polymorphism (SNP) arrays: either the UK BiLEVE Axiom array or the UK Biobank Axiom array. Variant imputation was performed using a joint Haplotype Reference Consortium and UK10,000 Genomes Project reference panel.

After excluding participants who had withdrawn consent to remain in the study, we additionally excluded participants whose genetic heterozygosity was beyond 10 standard deviations (SD) from the mean, along with participants whose genetic ancestry based on principal components (PC) 1 and 2 were beyond 10 SD of the mean of self-reported ‘White British’ participants. Any participant with a medical history of ocular surgery that potentially affected refractive error was also removed. Specifically, we considered: a history of cataract surgery (Data Field: 41200, C711-712, C718-719, C723, C751), corneal surgery (Data Field: 41200, C442, C444, C445, C448, C461-463, C465, C493), and self-reported refractive laser surgery (Data Field: 5325).

As reported^23^, a sample of 77,880 unrelated European-ancestry participants who had information available for both SER and AOSW were used as a vGWAS *training dataset* (Figure 1). Three vGWAS analyses – LT, CQR and SCAMPI – were carried out using this sample. All vGWAS analyses utilized the same set of approximately 1 million HapMap3 variants. Full details of the three vGWAS analyses are presented in the original publication^23^ and in Supplementary Note 1. A further 19,470 unrelated European-ancestry UKB participants with information for SER who were not included in the *training dataset* were used as the *validation dataset* (Table 1, Figure 1). An additional sample of n=221,516 UKB participants with known AOSW who were not used in the *training sample* or the *validation dataset* were used to form the *tuning sample* (Figure 1). A previously described formula^25^ was used to create a new phenotype “AOSW-inferred SER” for participants in the *tuning sample* based on age, sex, and AOSW (Supplementary Note 2).

**Figure 1.**
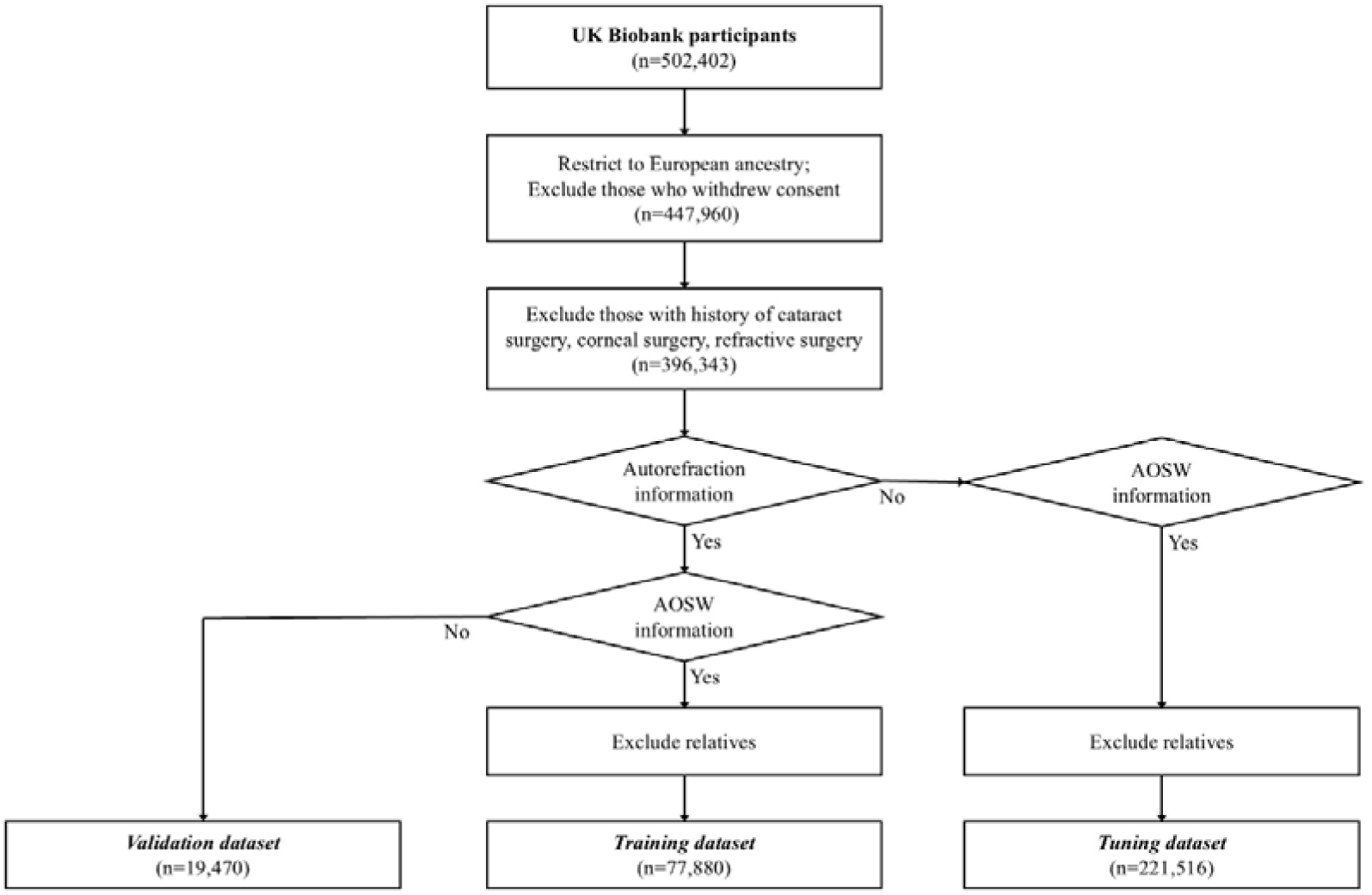
Selection scheme for UK Biobank participants.

**Table 1.** Demographic characteristics 538 of UK Biobank samples.

| **Characteristic** | ***Training dataset***<br>**(n=77,880)** | ***Tuning dataset***<br>**(n=221,516)** | ***Validation dataset***<br>**(n=19,470)** |
| --- | --- | --- | --- |
| Female (percentage) | 53.4% | 54.26% | 50.82% |
| Age (years) [mean ± SD] | 58.12 ± 7.54 | 57.21±7.56 | 55.15±8.79 |
| SER (D) [mean ± SD] | -0.30 ± 2.78 | NA | -0.12±2.16 |
| EduAge (years) [mean ± SD] | NA | 18.06±2.53 | 18.24±2.47 |
| AOSW (years) [median (Q1, Q3)] | 39 (15, 47) | 38 (15, 47) | NA |
| Abbreviations: SER: spherical equivalent refraction; EduAge: age of completed full-time education; AOSW: self-report age of onset of starting to wear spectacles or contact lenses; NA: not applicable. |  |  |  |

### ALSPAC cohort

The Avon longitudinal Study of Parents and Children (ALSPAC) cohort recruited pregnant women resident in Avon, UK with expected dates of delivery between 1st April 1991 and 31st December 1992. Of the initial pregnancies, 13,988 children were alive at 1 year of age. When the oldest children were approximately 7 years of age, an attempt was made to bolster the initial sample with eligible cases who had failed to join the study originally. This yielded a total sample size of 15,447. The health and wellbeing of participants was monitored through questionnaires and annual clinic visits.^26,27^ Ethical approval for the study was obtained from the ALSPAC Ethics and Law Committee and the Local Research Ethics Committees. Informed consent for the use of all data collected was obtained from participants following the recommendations of the ALSPAC Ethics and Law Committee at the time. Participants can contact the study team at any time to retrospectively withdraw consent for their data to be used. Study participation is voluntary and during all data collection sweeps, information was provided on the intended use of data. The study website contains details of all the data that is available through a fully searchable data dictionary and variable search tool: http://www.bristol.ac.uk/alspac/researchers/our-data/

Non-cycloplegic refractive error was measured when participants were aged approximately 15 years old. The SER of participants was calculated as described above for UKB participants. DNA samples from ALSPAC participants and their mothers were genotyped on the Illumina HumanHap660 quad chip genotyping platform and imputed using a 1000 Genomes Project reference panel.^26^ The time children spent reading and the time they spent outdoors at about 8 years old were ascertained from questionnaires answered by the parents or guardian, as described previously.^28^ For the time reading question, “How much time on average does your child spend reading books for pleasure on normal days in school holidays?”, responses of “Not at all” or “1 hour or less” were classified as “low” and coded as zero; while responses “1 to 2 hours” or “3 or more hours” were classified as “high” and coded as one. For the time outdoors question, “‘On a weekend day, how much time on average does your child spend each day out of doors in summer?’’, responses of “3 or more hours” were classified as “high” and coded as one; other responses were classified as “low” and coded as zero. There were 3471 children who had SER information at age 15 years, genotype data available, as well as information for time reading and time outdoors at age 8 years (Table 2).

**Table 2.** Demographic characteristics of ALSPAC samples at age of 15.

| **Characteristic** | **All**<br>**(n=3471)** | **Myopic**<br>**(n=1279)** | **Non-Myopic**<br>**(n=2192)** |
| --- | --- | --- | --- |
| Female (percentage) | 52.4% | 53.7% | 51.6% |
| Age [mean ± SD] | 15.41 ± 0.26 | 15.43 ± 0.28 | 15.42 ± 0.27 |
| SER [mean ± SD] | -0.39 ± 1.28 | -1.39 ± 1.24 | 0.19 ± 0.88 |
| Time spent reading<br>(low/high) at age of 8 years | 2118 / 1353<br>(61.0% / 39.0%) | 737 / 542<br>(57.6% / 42.4%) | 1381 / 811<br>(63.0% / 37.0%) |
| Time outdoors (low/high) at<br>age of 8 years | 315 / 3156<br>(9.1% / 90.9%) | 131 / 1148<br>(10.2% / 89.8%) | 184 / 2008<br>(8.4% / 91.6%) |
| Abbreviation: SER, spherical equivalent refraction. |  |  |  |

### vPGS construction and evaluation

Analyses were carried out using R v4.4.0. The vPGS evaluation scheme is shown in Figure 2. We applied two commonly used methodologies for re-weighting GWAS regression coefficients to account for linkage disequilibrium: pruning and thresholding (P+T) and LDpred2.^29,30^ For P+T, we tested ten different p-value thresholds using the PLINK v1.9 function --clump^31^: 5e-08, 1e-07, 1e-06, 1e-05, 1e-04, 1e-03, 1e-02, 0.05, 1e-01, and 1. For LDpred2, we tested an infinitesimal model as well as 40 different LDpred2-grid models: p-value thresholds of 1e-04, 1e-03, 1e-02, 1e-01 and 1, along with heritability values of 0.05, 0.1, 0.15 and 0.20, plus either a Full or Sparse model. vPGS were constructed for each of the three vGWAS methods: LT, CQR and SCAMPI. Single nucleotide polymorphism (SNP) effect sizes for vPGS construction were obtained as described in Supplementary Note 3. The optimal vPGS was selected as described below.

**Figure 2.**
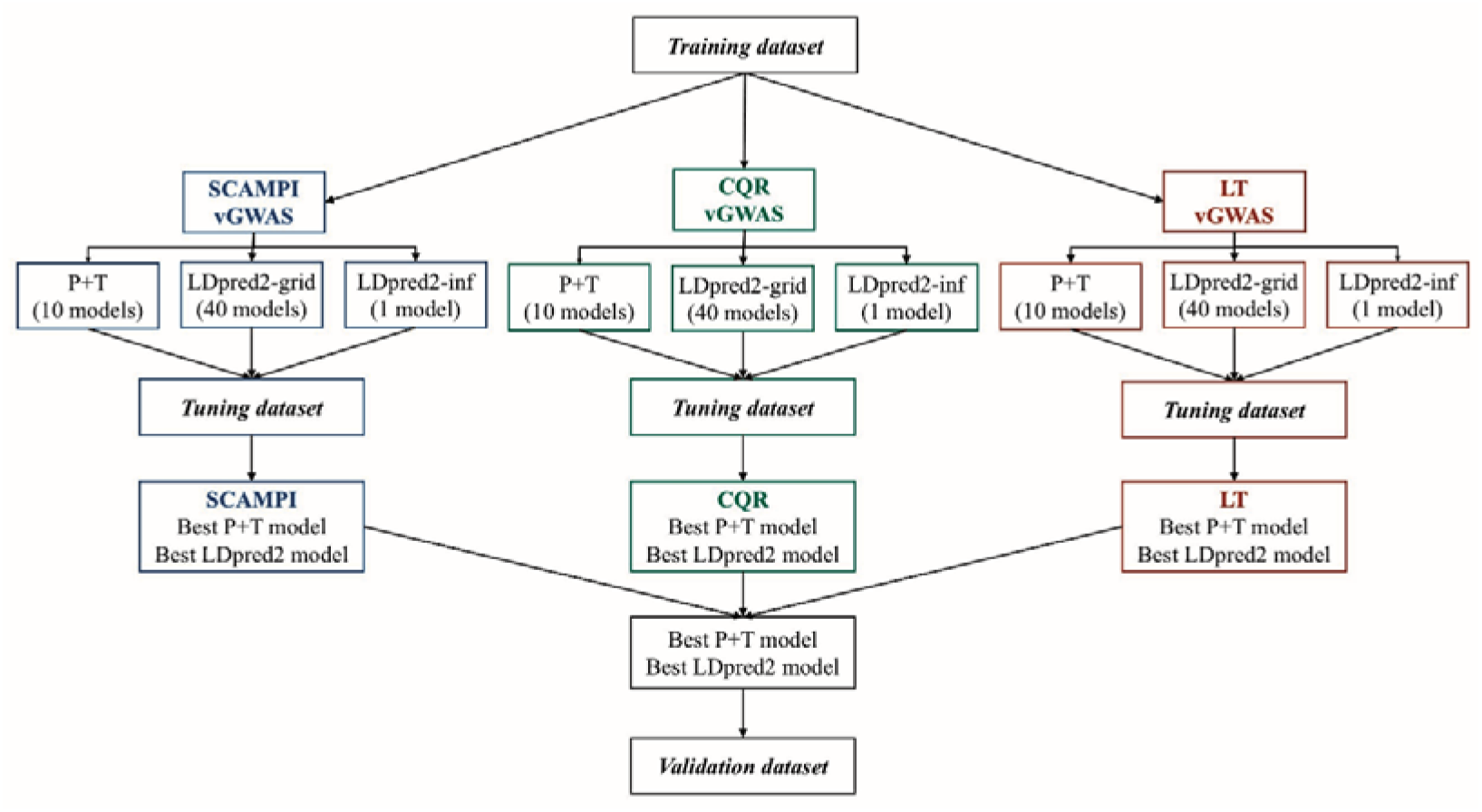
Schematic diagram of prioritization of vPGS models.

A conventional PGS was also constructed; this was done following a previously reported approach to construct a PGS for SER using data from UKB.^25^ A GWAS for SER was performed in the *training dataset* using BOLT-LMM,^32^ with age, squared age, sex, genotyping array and PC1-10 as covariates. Optimal LDpred2 parameter settings were chosen in a random sample of 4000 participants from the *validation dataset*. The accuracy of the conventional PGS was quantified as the incremental-R^2^ when comparing the following two linear regression models in the full *validation dataset*: Baseline model: AOSW-inferred SER ∼ age + age^2 + sex + genotyping array + PC1-10; Full model: AOSW-inferred SER ∼ age + age^2 + sex + genotyping array + PC1-10 + PGS.

Few studies have assessed vPGS performance, hence currently there is no consensus on how best to evaluate vPGS accuracy. Therefore, we adopted two metrics to quantify the phenotypic variance captured by a vPGS. The *tuning dataset* was divided into ten vPGS deciles (where decile one consisted of participants with the lowest vPGS values and decile ten participants with the highest vPGS values). Next, we fitted the following linear regression model in the *tuning dataset*: AOSW-inferred SER ∼ age + age^2 + sex + genotyping array + PC1-10 + mPGS (where mPGS is the optimal conventional PGS described in the paragraph above). The residuals from the regression model were used to calculate the variance of residual AOSW-inferred SER in each vPGS decile (Var_decile_). The first performance metric, ‘Diff’, was defined as: Diff = | Var_decile 10_ – Var_decile 1_ |. The second performance metric was the Spearman correlation between Var_decile_ and vPGS decile. Bootstrapping (with 10,000 iterations) was applied to acquire 95% confidence intervals (CI) for the Diff values and the Spearman correlation.

The six best-performing vPGS models from the *tuning dataset* – namely, the top P+T model and the top LDpred2 model for each of the three vGWAS methods – were taken forward for further evaluation in the *validation dataset* (n=19,470), using SER as the outcome phenotype. The capacity of the six best-performing vPGS models to detect variance in autorefraction-measured SER was assessed by calculating the Diff and Spearman correlation metrics in the *validation dataset*. The vPGS with the best performance in the *validation dataset* was taken forward as the optimal vPGS model. The optimal vPGS model and conventional PGS (mPGS) model have been made openly available at: https://doi.org/10.17035/cardiff.31963923

### Testing for vPGS × environmental risk factor interactions

The optimal vPGS model was used to test for a vPGS × EduAge and/or mPGS x EduAge interaction in the UKB *validation dataset*, using the following 3 equations:

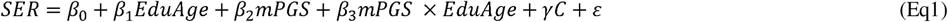

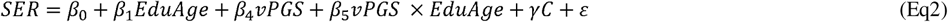

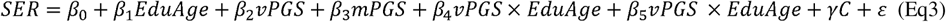

Where, in Eq3, β_0_ is an intercept term, β_1_, β_2_ and β_4_ are coefficients representing the main effects of EduAge, the conventional PGS (mPGS) and the vPGS, respectively, β_3_and β_5_ are mPGS-by-education and vPGS-by-education interaction terms; γ represents the coefficients of covariates (age, age^2, sex, genotyping array, and PC1-10), and ε is a normally distributed residual. *P* < 0.05 for the β_5_ term was considered evidence an association between education and SER that differed by vPGS.

In the ALSPAC sample (n=3471), we tested for a vPGS × time reading or a vPGS × time outdoors interaction using equivalent linear regression equations to Eq1-Eq3, except that EduAge was replaced by the binary variable for time reading or time outdoors. Taking advantage of the longitudinal nature of the ALSPAC study sample, these analyses assessed time reading and time outdoors when the participants were approximately age 8 years-old, but assessed SER when the participants were 15 years-old.

## Results

### Demographic characteristics of study cohorts

The selection of UKB participants is shown in Figure 1 and the demographic characteristics of the UKB *training dataset*, *tuning dataset* and *validation dataset* are shown in Table 1. The UKB participants were middle or older-aged adults who had completed their full-time education at 18 years of age, on average. The mean SER was close to emmetropia. Non-random selection of participants into the *training* and *validation datasets* led to an imbalance for most traits.

Specifically, participants in the *training dataset* were more likely to be female (*χ*^2^ = 147.13, *P* < 0.001), older (*t* = 43.43, *P* < 0.001), have a more negative SER (*t* =-9.63, *P* < 0.001), and to have spent more years in education (*t* = 2.72, *P* = 0.006). However, the magnitude of these differences was small (Table 1).

Participants from the longitudinal ALSPAC study who were included in the analysis sample were aged approximately 8 years-old when their time spent reading and spent outdoors was assessed, and 15 years-old when their (non-cycloplegic) SER was assessed (Table 2). The average SER was-0.39 D (SD 1.28). For time reading and time spent outdoors, which were coded as binary variables, 39.0% of ALSPAC participants spent a relatively “high” amount of time reading, while 90.9% spent a relatively “high” amount of time outdoors.

### Optimization of the vPGS model

Full results of the vPGS models in the *tuning dataset* are shown in Supplementary Tables S1-S3.

The Diff and Spearman correlation of the best-performing P+T and LDpred2 vPGS models for each of the three vGWAS methods in the *tuning dataset* are presented in Table 3. For the P+T models, the optimal p-value threshold for the SCAMPI, CQR and LT vGWAS summary statistics were *P* = 1E-04, *P* = 0.1 and *P* = 5E-08 respectively (Supplementary Figure). As regards the LDpred2 models, a total of 28, 40 and 30 models successfully converged for SCAMPI, CQR and LT, respectively. LDpred2-grid models consistently outperformed the LDpred2-infinitesimal model (Supplementary Tables S1-S3). For both SCAMPI and CQR, the optimal LDpred2-grid model was: *P* < 0.01; heritability = 0.05, sparsity = FALSE (Figure 3A,B). For LT, the optimal LDpred2-grid model was: *P* < 0.01; heritability = 0.05, sparsity = TRUE (Figure 3C). In general, the Diff and Spearman correlations were similar across a range of models and none of the three vGWAS methods stood out as a clearly superior method. Surprisingly, we observed that vPGS performance was dependent on sample size; all methods for generating vPGS tended to perform relatively well in the large *tuning dataset*, but when the best-performing P+T and LDpred2 vPGS models for each of the three vGWAS methods were evaluated in the much smaller *validation dataset* (n=19,470 vs. n=221,516), differences in vPGS performance were larger (Table 4; Figure 4). Finally, we selected the LDpred2-grid model from the CQR vGWAS as the top-performing vPGS model, based on the balance between its high Diff score yet competitive Spearman correlation (Table 4; Figure 4).

**Figure 3.**
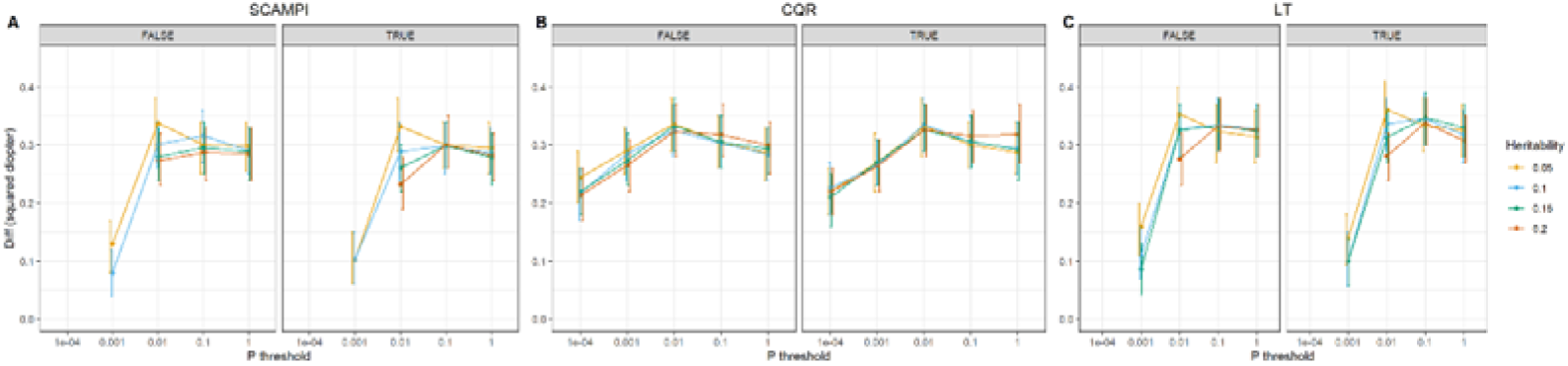
LDgrid2-grid model selection in the *tunning dataset*. Diff: the absolute difference between the variance of residual SER in 10^th^ vPGS decile vs. 1^st^ vPGS decile. **A**: vPGS constructed from SCAMPI summary statistics; **B**: vPGS constructed from CQR summary statistics; **C**: vPGS constructed from LT summary statistics. True and False refer to the sparsity parameter of the LDpred2 model. Results were obtained for the trait residuals of AOSW-inferred SER after adjusting for age, age^2, sex, genotyping array, PC1-10, and the optimal conventional PGS (mPGS).

**Figure 4.**
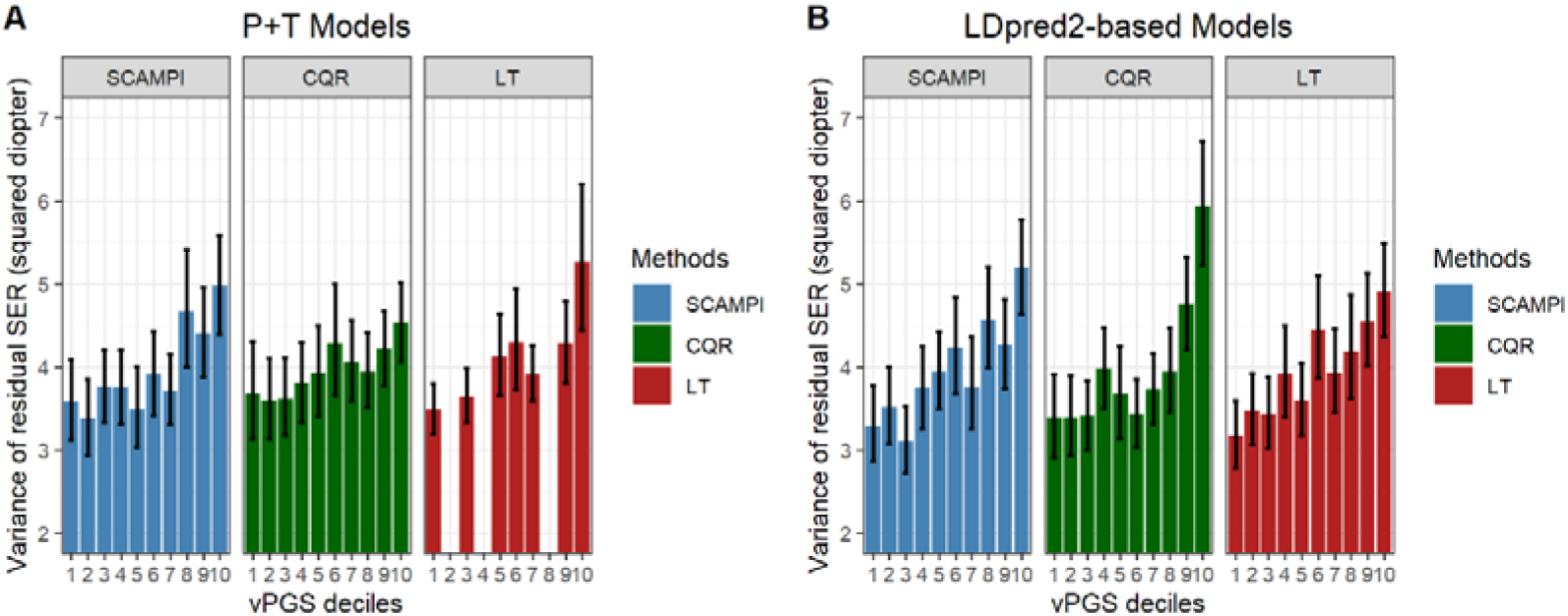
Spearman correlation of best-performing P+T and LDpred2-grid models. **A**: The three best-performing P+T models across vGWAS methods (rho = 0.78, 0.88 and 0.82, respectively); **B**: The three best-performing LDpred2-grid models across vGWAS methods (rho = 0.92, 0.87, 0.94, respectively). Results were obtained in the *validation dataset* for the trait residuals of SER after adjusting for age, age^2, sex, genotyping array, PC1-10, and the optimal conventional PGS (mPGS). Note the optimal P+T model for LT included only 3 variants; due to ties, this led to no participants being categorized into deciles 2, 4 or 8.

**Table 3.**
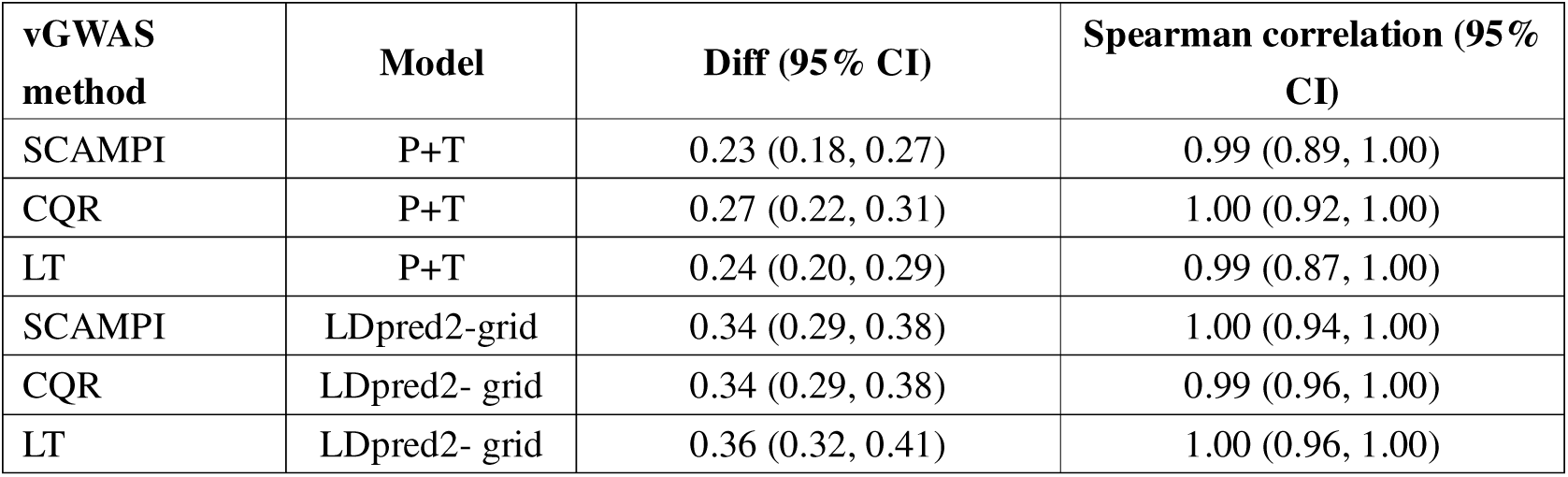
Summary of vPGS model optimization in the *tuning dataset*. Results are shown for the best-performing P+T model and the best-performing LDpred2 model for each of the three vGWAS methods. The test trait was residual AOSW-inferred SER after adjusting for age, age^2, sex, genotyping array, PC1-10, and the optimal conventional PGS (mPGS).

| **vGWAS method** | **Model** | **Diff (95% CI)** | **Spearman correlation (95% CI)** |
| --- | --- | --- | --- |
| SCAMPI | P+T | 0.23 (0.18, 0.27) | 0.99 (0.89, 1.00) |
| CQR | P+T | 0.27 (0.22, 0.31) | 1.00 (0.92, 1.00) |
| LT | P+T | 0.24 (0.20, 0.29) | 0.99 (0.87, 1.00) |
| SCAMPI | LDpred2-grid | 0.34 (0.29, 0.38) | 1.00 (0.94, 1.00) |
| CQR | LDpred2- grid | 0.34 (0.29, 0.38) | 0.99 (0.96, 1.00) |
| LT | LDpred2- grid | 0.36 (0.32, 0.41) | 1.00 (0.96, 1.00) |

**Table 4.** Summary of vPGS model optimization in the *validation dataset*. Results are shown for the models presented in Table 3. The test trait was residual SER after adjusting for age, age^2, sex, genotyping array, PC1-10, and the optimal conventional PGS (mPGS).

| vGWAS method | Model | Diff<br>(95% CI) | Spearman correlation<br>(95% CI) |
| --- | --- | --- | --- |
| SCAMPI | P+T | 1.40 (0.60, 2.21) | 0.78 (0.45, 0.93) |
| CQR | P+T | 0.85 (0.06, 1.59) | 0.88 (0.24, 0.93) |
| LT | P+T | 1.77 (0.89, 2.77) | 0.82 (0.36, 0.93) |
| SCAMPI | LDpred2-grid | 1.90 (1.20, 2.66) | 0.92 (0.61, 0.95) |
| CQR | LDpred2- grid | 2.55 (1.64, 3.47) | 0.87 (0.43, 0.93) |
| LT | LDpred2- grid | 1.73 (1.08, 2.46) | 0.94 (0.64, 0.96) |

### Performance of the conventional PGS

The optimal conventional PGS was obtained using the LDpred2 parameters: *P* < 0.01, heritability = 0.20, Sparsity = FALSE. In the independent *validation dataset*, the incremental R-squared for this conventional PGS was R^2^ = 0.09 (over-and-above the R-squared of a baseline model that included the covariates: age, age^2, sex, genotyping array, and PC1-10).

### Interactions between vPGS and environmental risk factors

We first tested for a vPGS × EduAge interaction in the UKB *validation dataset*. As well as evaluating a model with terms for a vPGS main effect and a vPGS × EduAge interaction, we also investigated models that included a conventional PGS (mPGS) main effect and an mPGS × EduAge interaction (either with or without the vPGS terms). As shown in Table 5, in all of these models, an additional year of full-time education (EduAge) was associated with a shift in SER of-0.12 D. In an analysis (model 2) that did not include the conventional PGS, a 1-SD increase in the optimal vPGS was associated with a shift in SER of-0.40 D (95% CI-0.64 to-0.16, *P* = 8.25E-04) as a main effect but there was no evidence of a vPGS × EduAge interaction (*P* = 0.76). Conversely, in an analysis (model 1) that did not include the vPGS, a 1-SD decrease in the conventional PGS, i.e. in the direction of greater susceptibility to myopia, was associated with a change in SER of-0.28 D (95%-0.04 to-0.52, *P* = 0.016) as well as a PGS × EduAge interaction of-0.02 D per SD decrease in PGS per year of additional education (95% CI-0.01 to-0.04, *P* = 2.86E-04). When both the conventional PGS and vPGS were included in the analysis model (model 3), the main effects of the PGS and vPGS reduced in magnitude (to-0.08 D per 1-SD decrease and-0.30 D per 1-SD increase, respectively). Furthermore, there was evidence of a PGS × EduAge interaction (-0.04 D per 1-SD decrease in PGS per year of education, 95% CI-0.02 to-0.05, *P* = 9.31E-06) as well as a vPGS × EduAge interaction (-0.02 D per 1-SD increase in vPGS per year of education, 95% CI-0.01 to-0.04, *P* = 0.011). In summary, these findings provided tentative evidence that the vPGS was associated with the risk of myopia in individuals undergoing extensive education, but that the magnitude of this association was much too small to be of clinical interest.

**Table 5.** Interaction between mPGS and vPGS and EduAge in the UKB validation dataset.

| Terms | Model 1 |  |  | Model 2 |  |  | Model 3 |  |  |
| --- | --- | --- | --- | --- | --- | --- | --- | --- | --- |
|  | BETA | SE | P | BETA | SE | P | BETA | SE | P |
| Intercept | 9.55 | 0.69 | 4.64E-43 | 10.17 | 0.72 | 1.41E-45 | 9.56 | 0.69 | 3.67E-43 |
| Sex (female) | 0.029 | 0.029 | 0.32 | 0.026 | 0.030 | 0.39 | 0.031 | 0.029 | 0.30 |
| Age (years) | -0.31 | 0.025 | 6.29E-35 | -0.33 | 0.026 | 1.77E-36 | -0.31 | 0.025 | 5.60E-35 |
| Squared age (years<sup>2</sup>) | 0.0031 | 0.0002 | 3.48E-41 | 0.0032 | 0.0002 | 4.12E-42 | 0.0031 | 0.0002 | 3.12E-41 |
| Genotyping array | -0.070 | 0.047 | 0.14 | -0.094 | 0.049 | 0.053 | -0.066 | 0.047 | 0.16 |
| PC1 | -0.004 | 0.009 | 0.62 | -0.0063 | 0.009 | 0.50 | -0.005 | 0.009 | 0.60 |
| PC2 | -0.0016 | 0.009 | 0.86 | -0.0037 | 0.010 | 0.70 | -0.0014 | 0.010 | 0.88 |
| PC3 | -0.0026 | 0.009 | 0.78 | -0.0015 | 0.010 | 0.88 | -0.0028 | 0.009 | 0.76 |
| PC4 | -0.0053 | 0.005 | 0.29 | -0.016 | 0.005 | 2.05E-03 | -0.0032 | 0.005 | 0.53 |
| PC5 | 0.0060 | 0.002 | 0.01 | -0.003 | 0.002 | 0.21 | 0.0078 | 0.002 | 9.00E-04 |
| PC6 | -0.0074 | 0.009 | 0.39 | -0.0057 | 0.009 | 0.51 | -0.008 | 0.009 | 0.35 |
| PC7 | -0.011 | 0.006 | 0.051 | -0.0092 | 0.006 | 0.12 | -0.012 | 0.006 | 0.044 |
| PC8 | -0.00012 | 0.006 | 0.98 | 0.0045 | 0.006 | 0.49 | -0.0006 | 0.006 | 0.92 |
| PC9 | -3.19E-05 | 0.004 | 0.99 | 0.0010 | 0.004 | 0.79 | 0.00033 | 0.004 | 0.93 |
| PC10 | 0.0094 | 0.007 | 0.18 | 0.0075 | 0.007 | 0.30 | 0.011 | 0.007 | 0.13 |
| **EduAge (years)** | **-0.12** | **0.006** | **5.79E-85** | **-0.12** | **0.006** | **1.64E-82** | **-0.12** | **0.006** | **3.57E-86** |
| **mPGS (Z-score)** | **0.28** | **0.12** | **1.60E-02** | - | - | - | **0.08** | **0.15** | **0.60** |
| **mPGS×EduAge** | **0.023** | **0.006** | **2.86E-04** | - | - | - | **0.037** | **0.008** | **9.31E-06** |
| **vPGS (Z-score)** | - | - | - | **-0.40** | **0.12** | **8.25E-04** | **-0.30** | **0.15** | **0.052** |
| **vPGS×EduAge** | - | - | - | **-0.002** | **0.006** | **0.76** | **0.021** | **0.008** | **0.011** |

Model 1. SER ~ EduAge + mPGS + mPGS × EduAge + covariates.
Model 2. SER ~ EduAge + vPGS + vPGS × EduAge + covariates.
Model 3. SER ~ EduAge + mPGS + mPGS × EduAge + vPGS + vPGS × EduAge + covariates.
Covariates: Sex, age, squared age, PC1-PC10, genotyping array.

Analogous models were fitted in the ALSPAC sample to investigate if either the vPGS × Time reading interaction or the vPGS × Time outdoors interaction was associated with SER (Tables 6 and 7). Across all models, spending a relatively high amount of time reading at age 8 years was associated with an SER at age 15 years that was-0.21 to-0.22 D more negative (myopic), as shown in Table 6. In the analysis that only considered the main effect and interaction effect of the vPGS (model 2), a 1-SD increase in the vPGS was associated with a similarly-sized shift towards myopia of-0.23 D (95% CI-0.28 to-0.18, *P* = 9.40E-18) but there was minimal evidence of a vPGS × Time reading interaction (*P* = 0.43). In the analysis that only considered the main effect and interaction effect of the conventional PGS (model 1), a 1-SD decrease in the PGS, i.e. in the direction of greater susceptibility to myopia, was associated with a larger-0.35 D shift towards myopia (95% CI 0.30 to 0.40, *P* = 1.40E-39), but again with no PGS × Time reading interaction (*P* = 0.15). In the analysis that included both the vPGS and the conventional PGS (model 3), the effect of the PGS remained similar (-0.35 D per 1-SD decrease, *P* = 3.89E-23) but the main effect of the vPGS was now non-significant (*P* = 0.89) and there remained negligible evidence of a PGS × Time reading interaction (*P* = 0.21) or a vPGS × Time reading interaction (*P* = 0.80).

**Table 6.** Interaction between mPGS and vPGS and Time Reading in the ALSPAC cohort.

| Terms | Model 1 |  |  | Model 2 |  |  | Model 3 |  |  |
| --- | --- | --- | --- | --- | --- | --- | --- | --- | --- |
|  | BETA | SE | P | BETA | SE | P | BETA | SE | P |
| Intercept | 47.12 | 34.30 | 0.17 | 50.98 | 35.22 | 0.15 | 46.97 | 34.31 | 0.17 |
| Sex (female) | 0.032 | 0.042 | 0.44 | 0.031 | 0.043 | 0.47 | 0.033 | 0.042 | 0.44 |
| Age (years) | -6.13 | 4.37 | 0.16 | -6.63 | 4.49 | 0.14 | -6.11 | 4.38 | 0.16 |
| Squared age (years<sup>2</sup>) | 0.20 | 0.14 | 0.15 | 0.22 | 0.14 | 0.13 | 0.20 | 0.14 | 0.16 |
| Time reading (high)<sup>¶</sup> | -0.21 | 0.043 | 1.29E-06 | -0.22 | 0.044 | 6.50E-07 | -0.21 | 0.043 | 1.25E-06 |
| PC1 | -22.26 | 11.36 | 0.05 | -19.36 | 11.67 | 0.10 | -22.49 | 11.37 | 0.048 |
| PC2 | 9.23 | 10.28 | 0.37 | 8.85 | 10.56 | 0.40 | 9.22 | 10.28 | 0.37 |
| PC3 | -8.48 | 9.23 | 0.36 | -10.83 | 9.48 | 0.25 | -8.44 | 9.23 | 0.36 |
| PC4 | -11.70 | 9.77 | 0.23 | -16.63 | 10.05 | 0.098 | -11.49 | 9.79 | 0.24 |
| PC5 | -5.94 | 9.90 | 0.55 | -3.86 | 10.16 | 0.70 | -5.86 | 9.90 | 0.55 |
| PC6 | -19.11 | 10.31 | 0.064 | -16.57 | 10.59 | 0.12 | -19.11 | 10.32 | 0.064 |
| PC7 | 0.073 | 10.03 | 0.99 | 5.90 | 10.29 | 0.57 | -0.016 | 10.03 | 0.99 |
| PC8 | -6.16 | 10.55 | 0.60 | -7.41 | 10.83 | 0.49 | -6.13 | 10.55 | 0.56 |
| PC9 | 12.57 | 10.06 | 0.21 | 14.69 | 10.33 | 0.16 | 12.50 | 10.06 | 0.21 |
| PC10 | -14.68 | 10.69 | 0.17 | -16.14 | 10.98 | 0.14 | -14.75 | 10.70 | 0.17 |
| mPGS (Z-score) | 0.35 | 0.026 | 1.40E-39 | - | - | - | 0.35 | 0.036 | 3.89E-23 |
| mPGS×Reading time | 0.061 | 0.043 | 0.15 | - | - | - | 0.071 | 0.057 | 0.21 |
| vPGS (Z-score) | - | - | - | -0.23 | 0.027 | 9.40E-18 | 0.0047 | 0.035 | 0.89 |
| vPGS×Reading time | - | - | - | -0.034 | 0.044 | 0.43 | 0.015 | 0.057 | 0.80 |

Model 1: SER ~ Time reading + mPGS + mPGS × Time reading + covariates.
Model 2: SER ~ Time reading + vPGS + vPGS × Time reading + covariates.
Model 3: SER ~ Time reading + mPGS + mPGS × Time reading + vPGS + vPGS × Time reading + covariates.
Covariates: Sex, age, squared age, PC1-PC10.
<sup>¶</sup>Time spent reading was assessed when participants were about 8 years old. SER and age were assessed when participants were about 15 years old.

**Table 7.** Interaction between mPGS and vPGS and Time Outdoors in the ALSPAC cohort.

| Terms | Model 1 |  |  | Model 2 |  |  | Model 3 |  |  |
| --- | --- | --- | --- | --- | --- | --- | --- | --- | --- |
|  | BETA | SE | P | BETA | SE | P | BETA | SE | P |
| Intercept | 46.98 | 34.40 | 0.17 | 49.67 | 35.34 | 0.16 | 46.72 | 34.42 | 0.17 |
| Sex (female) | -0.0022 | 0.042 | 0.96 | -0.0036 | 0.043 | 0.93 | -0.0021 | 0.042 | 0.96 |
| Age (years) | -6.11 | 4.39 | 0.16 | -6.47 | 4.51 | 0.15 | -6.08 | 4.39 | 0.17 |
| Squared age (years<sup>2</sup>) | 0.20 | 0.14 | 0.16 | 0.21 | 0.14 | 0.14 | 0.20 | 0.14 | 0.16 |
| Time outdoors (high)<sup>‡</sup> | 0.15 | 0.072 | 0.043 | 0.13 | 0.075 | 0.079 | 0.15 | 0.073 | 0.041 |
| PC1 | -20.43 | 11.39 | 0.073 | -17.99 | 11.70 | 0.12 | -20.61 | 11.40 | 0.071 |
| PC2 | 9.66 | 10.31 | 0.35 | 9.07 | 10.62 | 0.39 | 9.56 | 10.34 | 0.36 |
| PC3 | -9.69 | 9.25 | 0.29 | -12.01 | 9.50 | 0.21 | -9.69 | 9.26 | 0.30 |
| PC4 | -10.85 | 9.80 | 0.27 | -15.65 | 10.08 | 0.12 | -10.59 | 9.82 | 0.28 |
| PC5 | -6.58 | 9.93 | 0.51 | -4.58 | 10.20 | 0.65 | -6.59 | 9.93 | 0.51 |
| PC6 | -19.94 | 10.34 | 0.054 | -17.18 | 10.62 | 0.11 | -19.97 | 10.34 | 0.054 |
| PC7 | -0.51 | 10.06 | 0.96 | 5.41 | 10.33 | 0.60 | -0.63 | 10.06 | 0.95 |
| PC8 | -5.57 | 10.58 | 0.60 | -6.99 | 10.87 | 0.52 | -5.55 | 10.58 | 0.60 |
| PC9 | 11.2805 | 10.09 | 0.26 | 13.50 | 10.36 | 0.19 | 11.20 | 10.09 | 0.27 |
| PC10 | -13.67 | 10.73 | 0.20 | -15.44 | 11.03 | 0.16 | -13.70 | 10.74 | 0.20 |
| **mPGS (Z-score)** | 0.27 | 0.064 | 2.35E-05 | - | - | - | 0.29 | 0.089 | 1.35E-03 |
| **mPGS×Outdoor time** | 0.12 | 0.067 | 0.073 | - | - | - | 0.11 | 0.094 | 0.23 |
| **vPGS (Z-score)** | - | - | - | -0.19 | 0.070 | 6.5E-03 | 0.024 | 0.095 | 0.80 |
| **vPGS×Outdoor time** | - | - | - | -0.064 | 0.073 | 0.38 | -0.014 | 0.099 | 0.89 |

Model 1: SER ~ Time outdoors + mPGS + mPGS × Time outdoors + covariates.
Model 2: SER ~ Time outdoors + vPGS + vPGS × Time outdoors + covariates.
Model 3: SER ~ Time outdoors + mPGS + mPGS × Time outdoors + vPGS + vPGS × Time outdoors + covariates.
Covariates: Sex, age, squared age, PC1-PC10.
<sup>‡</sup>Time spent outdoors was assessed when participants were about 8 years old. SER and age were assessed when participants were about 15 years old.

Across all models testing for interactions with time outdoors (Table 7), the main effect of spending a relatively high amount of time outdoors at age 8 years was associated with an SER at age 15 years that was +0.13 to +0.15 D (*P* = 0.041 to 0.079) more positive (hyperopic). In the analysis examining just the main effect and interaction effect of the vPGS (model 2), a 1-SD increase in the vPGS was associated with a-0.19 D (95% CI-0.33 to-0.05, *P* = 6.5E-03) more negative SER, but there was minimal evidence for a vPGS × Time outdoors interaction (*P* = 0.38). In the analysis examining just the main effect and interaction effect of the conventional PGS (model 1), a 1-SD decrease in the PGS was associated with a-0.27 D (95% CI 0.14 to 0.40, *P* = 2.35E-05) more negative SER, again with scant evidence of a PGS × Time outdoors interaction (*P* = 0.07). In the analysis that included both the vPGS and the conventional PGS (model 3), the effect of the PGS remained similar (-0.29 D per 1-SD decrease, *P* = 1.35E-03) but the main effect of the vPGS was now non-significant (*P* = 0.80) and there was little evidence of a PGS × Time outdoors interaction (*P* = 0.23) or a vPGS × Time outdoors interaction (*P* = 0.89).

Thus, none of the analyses in the ALSPAC sample suggested that the vPGS was associated with the risk of myopia preferentially in individuals with high levels of reading or low levels of time outdoors.

## Discussion

This study investigated if a vPGS could be used as a tool to identify children at high risk of myopia via exposure to lifestyle risk factors such as excessive near work or insufficient time outdoors. We evaluated six methods for constructing a vPGS (three vGWAS protocols × two variant reweighting schemes), which covered both established and newly-reported approaches for identifying vQTL. The optimal vPGS was selected based on two performance metrics: Diff and Spearman correlation. When participants were grouped into deciles of the optimal vPGS (and after adjusting for a conventional PGS for SER), the variance of SER in the first vs. last vPGS decile had non-overlapping 95% CIs, demonstrating that the optimal vPGS could stratify participants into groups with relatively low or high variance (Figure 4). However, there were fluctuations in SER variance across vPGS deciles (Figure 4), implying that the prediction of SER variance by the optimal vPGS was noisy in comparison to the prediction of SER by a conventional PGS. Although there was tentative evidence of a vPGS × education interaction in an independent sample of UKB participants, the effect size was very small (-0.02 D per 1-SD increase in vPGS per year of additional education). Furthermore, in a longitudinal study of children aged 8–15 years-old, there was minimal evidence of a vPGS × time spent reading interaction or a vPGS × time spent outdoors interaction, suggesting that the vPGS did not show promise as a clinical tool for identifying children at high risk of myopia via exposure to lifestyle risk factors.

Few prior studies have investigated the capacity of a vPGS to identify individuals with high “phenotypic plasticity” who may have enhanced susceptibility to gene-environment interactions. Therefore, the criteria used to evaluate vPGS performance are not well established^19,20^. Miao and colleagues ^20^ investigated the ability of vPGS to capture the variance of BMI in three cohorts; BMI variance was 52% to 73% higher in the first vs. last vPGS quintile (although fluctuations in BMI variance across vPGS quintiles was evident, as we found for our vPGS for SER). Miao et al.^20^ quantified population-level variability of vPGS based on the concept of “integrated rank scores” developed in their study. Johnson et al.^19^ regressed BMI on covariates and regarded the squared residuals as a proxy of population-level phenotypic variability to gauge vPGS accuracy. Thus, defining a metric that quantifies the phenotypic variance of an individual person is a thorny concept. In the current study, we quantified vPGS accuracy using two parameters relating to vPGS deciles. First, the Diff value was used to assess the difference in variance captured by the vPGS in the first vs. last decile. Second, the Spearman correlation between the phenotypic variance of each vPGS decile vs. decile number was used to assess if the variance captured by a vPGS followed a consistent trend. Based on Diff value and the Spearman correlation, all three vGWAS methods yielded promising vPGS that were capable of stratifying a population into deciles with relatively high or low SER variance. The optimal vPGS, which was derived using LDpred2-grid reweighting of CQR vGWAS summary statistics, was able to stratify participants such that the SER variance of individuals in the tenth vPGS decile was 75% greater than that of participants in the first vPGS decile. Thus, the performance of this CQR-based vPGS for SER was on a par with the vPGS for BMI reported by Miao et al.^20^

Subsequent evaluation of the optimum vPGS suggested it had little or no capacity to identify individuals who were at an above-average genetic risk of developing myopia through exposure to lifestyle risk factors such as education, reading, or insufficient time outdoors (Tables 5-7). From these findings, we cautiously infer that genetic susceptibility to myopia is primarily conferred directly (i.e. by conventional PGS × lifestyle risk factor interactions) rather than indirectly through modified phenotypic plasticity (i.e. vPGS × lifestyle risk factor interactions).

To our knowledge, only four previous studies have explored vPGS × environment interactions^19–22^. Two of these studies^20,22^ constructed vPGS that included all independent vGWAS variants (equivalent to clumping with a p-value threshold of 1). Specifically, Miao et al.^20^ used a quantile regression vGWAS method (QUAIL) to construct a vPGS and found significant vPGS × physical activity/sedentary behavior interactions associated with BMI. Xiang et al.^22^ applied the same vPGS construction method to Levene’s test vGWAS summary statistics for several blood cell traits. They found that individuals in the top 5% vs. bottom 5% of the blood cell trait vPGS had a higher alcohol intake (subsequent Mendelian randomization analysis further supported a causal relationship between alcohol intake and blood cell trait variability).^22^ By including all non-imputed vGWAS variants without clumping, Johnson et al.^19^ built a series of vPGS for traits such as education level, BMI and height. They tested for vPGS × environment interactions in relation to the societal effect of an educational reform, using an instrumental variable framework. Schmitz et al.^21^ also constructed a vPGS for BMI using all available independent variants from a vGWAS; they studied the effect of job loss on BMI and found its effects were moderated by BMI plasticity (as indexed by the vPGS). As regards SER, our results suggested that including all independent variants in a vGWAS was not the optimal method for constructing a vPGS. Instead, we found that LDpred2-grid provided optimal performance (Figure 3), although the performance gain was more modest for a vPGS than is typically found for a PGS.^30^

Unlike most prior studies, we adjusted for the effects of a conventional PGS when assessing vPGS accuracy. Such adjustment is potentially advantageous because the association of genetic variants with traits leads to a mean-variance relationship.^33^ Thus, a conventional PGS that is designed to detect the mean effects on a trait of a set of genetic variants is also expected to be associated with the variance of the trait. Likewise, a vPGS is expected to be associated with the mean level of the trait as well as its variance (this was indeed the case in the current study; the optimal vPGS was associated with SER level as well as with SER variance; Tables 5-7). The mean-variance relationship typically induces a correlation of PGS vs. vPGS scores for a study sample. Therefore, to demonstrate that a vPGS provides information that is distinct from that already provided by a conventional PGS, it is necessary to account for the effects of the PGS and vPGS simultaneously. Accordingly, we included both a conventional PGS and a vPGS in models testing for vPGS × lifestyle risk factor interactions associated with SER.

In summary, we constructed a vPGS for SER that was capable of stratifying an independent sample into groups with relatively high or low SER variance. Despite the theoretical appeal of using a vPGS to quantify a person’s “phenotypic plasticity”, we found that exposure to lifestyle risk factors such as education, reading, or insufficient time outdoors was associated with similar changes in SER in individuals with high or low vPGS values. Since previous work has demonstrated that genetic susceptibility to myopia indexed by a conventional PGS predicts the shift in SER associated with an additional year of education^17^, we conclude that – based on currently available methods and datasets – a conventional PGS for SER outperforms a vPGS in identifying children at above-average risk of myopia from exposure to lifestyle risk factors.

## Acknowledgements

UK Biobank was established by the Wellcome Trust; the UK Medical Research Council; the Department for Health (London, UK); Scottish Government (Edinburgh, UK); and the Northwest Regional Development Agency (Warrington, UK). It also received funding from the Welsh Assembly Government (Cardiff, UK); the British Heart Foundation; and Diabetes UK. Collection of eye and vision data was supported by The Department for Health through an award made by the NIHR to the Biomedical Research Centre at Moorfields Eye Hospital NHS Foundation Trust, and UCL Institute of Ophthalmology, London, United Kingdom (grant no.

BRC2_009). Additional support was provided by The Special Trustees of Moorfields Eye Hospital, London, United Kingdom (grant no. ST 12 09). This research was conducted using the UK Biobank Resource under Application Number 83325. <u>ALSPAC</u>: We are extremely grateful to all the families who took part in this study, the midwives for their help in recruiting them, and the whole ALSPAC team, which includes data collection staff, data and administrations staff, technical managers and the technical staff with the Bristol Bioresource Laboratory, based within the University of Bristol. Data analysis was performed on the HAWK computing cluster, managed by Supercomputing Wales and Cardiff University ARCCA.

## Data availability statement

UK Biobank data are available via application at https://www.ukbiobank.ac.uk. vGWAS summary statistics generated in this study for SCAMPI, Levene’s Test and CQR have been deposited at: https://doi.org/10.17035/cardiff.30814814. Summary statistics for the optimal vPGS and mPGS constructed in this study have been deposited at: https://doi.org/10.17035/cardiff.31963923.

## Author contribution

XH and JAG conceived and designed the project. LT and JG provided supervision and arranged access to datasets. XH performed the data analysis. All authors contributed to interpretation of data analysis results. XH and JG had access to the raw data and verified the data. XH wrote and prepared the Article. All authors had access to the data presented, read, revised and approved the final article.

