## Supplementary Material for "Variance polygenic scores (vPGS) as a tool for studying gene-environment interactions associated with refractive error"

### Supplementary Information

### Supplementary Note 1. vGWAS analysis details.

As described previously[1], we applied three methods to conduct vGWAS analyses: SCAMPI (Scalable Cauchy Aggregate test using Multiple Phenotypes to test Interactions),[2] conditional quantile regression (CQR),[3] and Levene's test (LT).[4] Based on SCAMPI's requirement to have multiple phenotypes as input, we selected an analysis sample of n=77,880 UK Biobank participants with information available for both SER and AOSW (Figure 1; *training dataset*). All participants were unrelated, had European genetic ancestry, and did not have a history of past surgical treatment likely to affect SER (see Methods).

The SER and AOSW phenotypes were each regressed on the covariates, age, age^2, sex, genotyping array, and PC1-10. Phenotypes were not transformed to a normal distribution. Approximately 1 million HapMap3 variants (https://doi.org/10.5281/zenodo.7773502) were included in each vGWAS analysis. The R package SCAMPI (https://github.com/epstein-software/SCAMPI) was run using the default settings for the combined residual SER and residual AOSW phenotypes. SCAMPI outputs several p-values: here, we used the p-value for the SER phenotype. CQR was performed for the residual SER phenotype, using the rq function from the R package quantreg version 6.1, at nine quantile values (0.1, 0.2, 0.3, … 0.9). SNP regression coefficients ($\beta_{SNP}$) at the nine quantiles ($\tau$) were fit with an inverse variance-weighted restricted maximum likelihood meta-regression model that included an intercept term, a linear term ($\beta_{SNP} \times \tau$) and a quadratic term ($\beta_{SNP} \times \tau^2$) using the rma function from the R package metafor version 4.8-0. Here, we used the CQR meta-regression quadratic term to build the vPGS. Levene's median test was performed with OSCA (--vqtl option),[4] using the residual SER phenotype. OSCA outputs both a beta coefficient and a p-value for Levene's test.

### Supplementary Note 2. Derivation of AOSW-inferred SER phenotype.

The AOSW-inferred phenotype has been used in previous studies.[5,6] Here, the *training dataset* (n = 77,880), which contained information about participants' SER and AOSW, was used to train a predictive model for "AOSW-inferred SER". Five-fold cross-validation was used for optimizing the parameters of predictors in the following equation:

$$\mathrm{AOSW-inferred\ SER} \sim \mathrm{sex} + \mathrm{poly(AOSW, n)} + \mathrm{poly(age, m)}$$

Where, poly($a$, $b$) is a R function to generate orthogonal polynomials up to order $b$ for predictor $a$. The parameter, $b$ was tested across the range m = 1–6 for age, and n = 1–15 for AOSW. Based on the Akaike Information Criterion (AIC) of the 5-fold cross-validation analysis, optimal settings were, n = 13 and m = 6. This model was used to predict the AOSW-inferred SER of participants in the *tuning dataset* (n = 221,516).

### Supplementary Note 3. vGWAS effect sizes for vPGS construction.

Beta coefficients quantifying SNP effect sizes are required to construct a PGS or vPGS. SNP effect sizes for the CQR vGWAS meta-regression quadratic term were used as the effect size for building a CQR vPGS. The beta coefficients from the OSCA Levene's median test were used as the effect size for LT vPGS construction. SCAMPI outputs p-values but not beta coefficients. Therefore, the p-values form SCAMPI were transformed to Z-values (formula 1), where Φ is the standard normal cumulative distribution function. The sign of the Z-value for each SNP was borrowed from the Levene's test vGWAS summary statistics. The standard error (SE) of the effect size of each variant was calculated (formula 2), which required the sample size (N) and minor allele frequencies (MAF) of each variant. Finally, the SNP effect size (β) was calculated (formula 3).

$$|Z| = \Phi^{-1}\left(1 - \frac{P}{2}\right) \qquad (1)$$

$$SE = \frac{1}{\sqrt{2N \times MAF \times (1 - MAF)}} \qquad (2)$$

$$\beta = Z \times SE \qquad (3)$$

**Supplementary Table S1. Performance of SCAMPI vPGS models in the *tuning* *dataset*.**

| Method | Tuning parameters | Diff (95% CI) | Spearman Correlation (95% CI) |
|---|---|---|---|
| P+T | P<5.00E-08 | 0.18 (0.13, 0.22) | 1.00 (0.87, 1.00) |
| P+T | P<1.00E-07 | 0.20 (0.15, 0.24) | 0.99 (0.90, 1.00) |
| P+T | P<1.00E-06 | 0.21 (0.17, 0.26) | 0.96 (0.89, 1.00) |
| P+T | P<1.00E-05 | 0.18 (0.14, 0.23) | 0.98 (0.85, 0.99) |
| P+T | P<1.00E-04 | 0.23 (0.18, 0.27) | 0.99 (0.89, 1.00) |
| P+T | P<1.00E-03 | 0.23 (0.18, 0.27) | 0.98 (0.83, 0.99) |
| P+T | P<1.00E-02 | 0.20 (0.15, 0.24) | 0.99 (0.84, 0.99) |
| P+T | P<5.00E-02 | 0.18 (0.14, 0.23) | 1.00 (0.90, 1.00) |
| P+T | P<1.00E-01 | 0.20 (0.16, 0.24) | 0.96 (0.89, 0.99) |
| P+T | P<1.00 | 0.20 (0.16, 0.25) | 0.99 (0.89, 1.00) |
| LDpred2-inf | - | 0.29 (0.24, 0.33) | 1.00 (0.90, 1.00) |
| LDpred2-grid | P=0.001; $H^2$=0.05; S=FALSE | 0.13 (0.08, 0.17) | 0.94 (0.71, 0.95) |
| LDpred2-grid | P=0.01; $H^2$=0.05; S=FALSE | 0.34 (0.29, 0.38) | 1.00 (0.94, 1.00) |
| LDpred2-grid | P=0.1 $H^2$=0.05; S=FALSE | 0.30 (0.25, 0.34) | 1.00 (0.93, 1.00) |
| LDpred2-grid | P=1; $H^2$=0.05; S=FALSE | 0.30 (0.26, 0.34) | 1.00 (0.92, 1.00) |
| LDpred2-grid | P=0.001; $H^2$=0.1; S=FALSE | 0.08 (0.04, 0.12) | 0.94 (0.56, 0.95) |
| LDpred2-grid | P=0.01; $H^2$=0.1; S=FALSE | 0.30 (0.26, 0.34) | 1.00 (0.95, 1.00) |
| LDpred2-grid | P=0.1 $H^2$=0.1; S=FALSE | 0.32 (0.27, 0.36) | 0.99 (0.94, 1.00) |
| LDpred2-grid | P=1; $H^2$=0.1; S=FALSE | 0.29 (0.25, 0.33) | 0.95 (0.89, 1.00) |
| LDpred2-grid | P=0.01; $H^2$=0.15; S=FALSE | 0.28 (0.24, 0.33) | 1.00 (0.93, 1.00) |
| LDpred2-grid | P=0.1; $H^2$=0.15; S=FALSE | 0.30 (0.25, 0.34) | 0.99 (0.93, 1.00) |
| LDpred2-grid | P=1 $H^2$=0.15; S=FALSE | 0.29 (0.24, 0.33) | 0.99 (0.91, 1.00) |
| LDpred2-grid | P=0.01; $H^2$=0.20; S=FALSE | 0.27 (0.23, 0.32) | 0.98 (0.89, 1.00) |
| LDpred2-grid | P=0.1; $H^2$=0.20; S=FALSE | 0.29 (0.24, 0.33) | 0.99 (0.92, 1.00) |
| LDpred2-grid | P=1 $H^2$=0.20; S=FALSE | 0.29 (0.24, 0.33) | 0.99 (0.89, 0.99) |
| LDpred2-grid | P=0.001; $H^2$=0.05; S=TRUE | 0.10 (0.06, 0.15) | 0.87 (0.61, 0.96) |
| LDpred2-grid | P=0.01; $H^2$=0.05; S=TRUE | 0.33 (0.29, 0.38) | 1.00 (0.93, 1.00) |
| LDpred2-grid | P=0.1 $H^2$=0.05; S=TRUE | 0.30 (0.26, 0.34) | 0.99 (0.94, 1.00) |
| LDpred2-grid | P=1; $H^2$=0.05; S=TRUE | 0.30 (0.25, 0.34) | 1.00 (0.94, 1.00) |
| LDpred2-grid | P=0.001; $H^2$=0.10; S=TRUE | 0.10 (0.06, 0.15) | 0.94 (0.61, 0.95) |
| LDpred2-grid | P=0.01; $H^2$=0.10; S=TRUE | 0.29 (0.24, 0.34) | 0.99 (0.92, 1.00) |
| LDpred2-grid | P=0.1 $H^2$=0.10; S=TRUE | 0.30 (0.25, 0.34) | 0.98 (0.92, 1.00) |
| LDpred2-grid | P=1; $H^2$=0.10; S=TRUE | 0.29 (0.25, 0.33) | 0.99 (0.90, 1.00) |
| LDpred2-grid | P=0.01; $H^2$=0.15; S=TRUE | 0.26 (0.22, 0.30) | 1.00 (0.93, 1.00) |
| LDpred2-grid | P=0.1 $H^2$=0.15; S=TRUE | 0.30 (0.26, 0.34) | 1.00 (0.95, 1.00) |
| LDpred2-grid | P=1; $H^2$=0.15; S=TRUE | 0.28 (0.23, 0.32) | 1.00 (0.90, 1.00) |
| LDpred2-grid | P=0.01; $H^2$=0.20; S=TRUE | 0.23 (0.19, 0.28) | 0.99 (0.89, 1.00) |
| LDpred2-grid | P=0.1 $H^2$=0.20; S=TRUE | 0.30 (0.26, 0.35) | 0.99 (0.94, 1.00) |
| LDpred2-grid | P=1; $H^2$=0.20; S=TRUE | 0.28 (0.24, 0.32) | 1.00 (0.92, 1.00) |

P=P threshold; $H^2$=heritability; S=sparsity.

**Supplementary Table S2. Performance of CQR vPGS models in the *tuning* *dataset*.**

| Method | Tuning parameters | Diff (95% CI) | Spearman Correlation (95% CI) |
|---|---|---|---|
| P+T | P<5.00E-08 | 0.16 (0.11, 0.20) | 0.96 (0.85, 0.96) |
| P+T | P<1.00E-07 | 0.16 (0.11, 0.20) | 0.96 (0.85, 0.96) |
| P+T | P<1.00E-06 | 0.15 (0.10, 0.19) | 0.99 (0.83, 0.99) |
| P+T | P<1.00E-05 | 0.18 (0.14, 0.23) | 0.96 (0.82, 0.99) |
| P+T | P<1.00E-04 | 0.18 (0.13, 0.22) | 0.90 (0.77, 0.99) |
| P+T | P<1.00E-03 | 0.24 (0.20, 0.29) | 0.98 (0.87, 0.99) |
| P+T | P<1.00E-02 | 0.22 (0.17, 0.26) | 1.00 (0.88, 1.00) |
| P+T | P<5.00E-02 | 0.24 (0.20, 0.28) | 0.96 (0.90, 1.00) |
| P+T | P<1.00E-01 | 0.27 (0.22, 0.31) | 1.00 (0.92, 1.00) |
| P+T | P<1.00 | 0.26 (0.22, 0.30) | 0.99 (0.93, 1.00) |
| LDpred2-inf | - | 0.283 (0.24, 0.33) | 0.99 (0.94, 1.00) |
| LDpred2-grid | P=0.0001; $H^2$=0.05; S=FALSE | 0.24 (0.2, 0.29) | 1.00 (0.89, 1.00) |
| LDpred2-grid | P=0.001; $H^2$=0.05; S=FALSE | 0.29 (0.25, 0.33) | 0.99 (0.93, 0.99) |
| LDpred2-grid | P=0.01; $H^2$=0.05; S=FALSE | 0.34 (0.29, 0.38) | 0.99 (0.96, 1.00) |
| LDpred2-grid | P=0.1 $H^2$=0.05; S=FALSE | 0.30 (0.26, 0.35) | 0.99 (0.93, 1.00) |
| LDpred2-grid | P=1; $H^2$=0.05; S=FALSE | 0.29 (0.24, 0.33) | 0.99 (0.94, 1.00) |
| LDpred2-grid | P=0.0001; $H^2$=0.1; S=FALSE | 0.22 (0.17, 0.26) | 0.99 (0.89, 1.00) |
| LDpred2-grid | P=0.001; $H^2$=0.1; S=FALSE | 0.29 (0.24, 0.33) | 0.99 (0.93, 1.00) |
| LDpred2-grid | P=0.01; $H^2$=0.1; S=FALSE | 0.33 (0.28, 0.37) | 0.98 (0.94, 0.99) |
| LDpred2-grid | P=0.1 $H^2$=0.1; S=FALSE | 0.30 (0.26, 0.35) | 1.00 (0.95, 1.00) |
| LDpred2-grid | P=1; $H^2$=0.1; S=FALSE | 0.28 (0.24, 0.33) | 0.99 (0.94, 1.00) |
| LDpred2-grid | P=0.0001; $H^2$=0.15; S=FALSE | 0.22 (0.18, 0.26) | 0.99 (0.90, 1.00) |
| LDpred2-grid | P=0.001; $H^2$=0.15; S=FALSE | 0.27 (0.23, 0.32) | 0.96 (0.91, 0.99) |
| LDpred2-grid | P=0.01; $H^2$=0.15; S=FALSE | 0.33 (0.29, 0.38) | 0.99 (0.95, 1.00) |
| LDpred2-grid | P=0.1; $H^2$=0.15; S=FALSE | 0.31 (0.26, 0.35) | 0.99 (0.93, 1.00) |
| LDpred2-grid | P=1 $H^2$=0.15; S=FALSE | 0.29 (0.25, 0.33) | 0.99 (0.93, 0.99) |
| LDpred2-grid | P=0.0001; $H^2$=0.20; S=FALSE | 0.21 (0.17, 0.26) | 0.96 (0.89, 0.99) |
| LDpred2-grid | P=0.001; $H^2$=0.20; S=FALSE | 0.27 (0.22, 0.31) | 0.96 (0.93, 1.00) |
| LDpred2-grid | P=0.01; $H^2$=0.20; S=FALSE | 0.32 (0.28, 0.37) | 0.99 (0.93, 1.00) |
| LDpred2-grid | P=0.1; $H^2$=0.20; S=FALSE | 0.32 (0.28, 0.37) | 0.99 (0.94, 1.00) |
| LDpred2-grid | P=1 $H^2$=0.20; S=FALSE | 0.30 (0.25, 0.34) | 0.96 (0.95, 1.00) |
| LDpred2-grid | P=0.0001; $H^2$=0.05; S=TRUE | 0.22 (0.18, 0.26) | 0.99 (0.90, 1.00) |
| LDpred2-grid | P=0.001; $H^2$=0.05; S=TRUE | 0.27 (0.22, 0.32) | 0.98 (0.92, 0.99) |
| LDpred2-grid | P=0.01; $H^2$=0.05; S=TRUE | 0.33 (0.28, 0.38) | 0.98 (0.94, 1.00) |
| LDpred2-grid | P=0.1 $H^2$=0.05; S=TRUE | 0.30 (0.26, 0.34) | 0.99 (0.93, 1.00) |
| LDpred2-grid | P=1; $H^2$=0.05; S=TRUE | 0.29 (0.25, 0.34) | 0.99 (0.94, 1.00) |
| LDpred2-grid | P=0.0001; $H^2$=0.10; S=TRUE | 0.23 (0.18, 0.27) | 0.99 (0.88, 0.99) |
| LDpred2-grid | P=0.001; $H^2$=0.10; S=TRUE | 0.27 (0.23, 0.31) | 0.98 (0.92, 1.00) |
| LDpred2-grid | P=0.01; $H^2$=0.10; S=TRUE | 0.34 (0.29, 0.38) | 0.98 (0.95, 1.00) |
| LDpred2-grid | P=0.1 $H^2$=0.10; S=TRUE | 0.31 (0.26, 0.35) | 0.99 (0.94, 1.00) |
| LDpred2-grid | P=1; $H^2$=0.10; S=TRUE | 0.29 (0.25, 0.34) | 0.99 (0.94, 1.00) |
| LDpred2-grid | P=0.0001; $H^2$=0.15; S=TRUE | 0.21 (0.16, 0.25) | 0.98 (0.89, 1.00) |
| LDpred2-grid | P=0.001; $H^2$=0.15; S=TRUE | 0.27 (0.23, 0.31) | 1.00 (0.93, 1.00) |
| LDpred2-grid | P=0.01; $H^2$=0.15; S=TRUE | 0.33 (0.29, 0.37) | 0.98 (0.95, 1.00) |
| LDpred2-grid | P=0.1 $H^2$=0.15; S=TRUE | 0.31 (0.26, 0.35) | 0.96 (0.94, 1.00) |
| LDpred2-grid | P=1; $H^2$=0.15; S=TRUE | 0.29 (0.24, 0.34) | 1.00 (0.94, 1.00) |
| LDpred2-grid | P=0.0001; $H^2$=0.20; S=TRUE | 0.22 (0.18, 0.26) | 0.95 (0.88, 0.99) |
| LDpred2-grid | P=0.001; $H^2$=0.20; S=TRUE | 0.26 (0.22, 0.31) | 0.96 (0.90, 0.99) |
| LDpred2-grid | P=0.01; $H^2$=0.20; S=TRUE | 0.33 (0.28, 0.37) | 1.00 (0.95, 1.00) |
| LDpred2-grid | P=0.1 $H^2$=0.20; S=TRUE | 0.32 (0.27, 0.36) | 0.99 (0.95, 1.00) |
| LDpred2-grid | P=1; $H^2$=0.20; S=TRUE | 0.32 (0.27, 0.37) | 0.99 (0.95, 1.00) |

P=P threshold for LDpred2-grid model; $H^2$=heritability; S=sparsity.

**Supplementary Table S3. Performance of LT vPGS models in the *tuning dataset*.**

| Method | Tuning parameters | Diff (95% CI) | Spearman Correlation (95% CI) |
|---|---|---|---|
| P+T | P<5.00E-08 | 0.24 (0.19, 0.28) | 0.93 (0.81, 0.99) |
| P+T | P<1.00E-07 | 0.24 (0.19, 0.28) | 0.93 (0.83, 0.99) |
| P+T | P<1.00E-06 | 0.23 (0.18, 0.27) | 0.99 (0.85, 1.00) |
| P+T | P<1.00E-05 | 0.20 (0.15, 0.24) | 0.98 (0.87, 1.00) |
| P+T | P<1.00E-04 | 0.15 (0.11, 0.20) | 0.94 (0.84, 0.99) |
| P+T | P<1.00E-03 | 0.17 (0.13, 0.22) | 0.96 (0.82, 0.99) |
| P+T | P<1.00E-02 | 0.22 (0.18, 0.27) | 0.98 (0.84, 0.99) |
| P+T | P<5.00E-02 | 0.24 (0.20, 0.29) | 0.99 (0.83, 1.00) |
| P+T | P<1.00E-01 | 0.24 (0.20, 0.29) | 0.99 (0.87, 1.00) |
| P+T | P<1.00 | 0.24 (0.20, 0.29) | 0.99 (0.81, 0.99) |
| LDpred2-infinitesimal | - | 0.32 (0.27, 0.36) | 0.99 (0.94, 1.00) |
| LDpred2-grid | P=0.001; $H^2$=0.05; S=FALSE | 0.16 (0.11, 0.20) | 0.99 (0.61, 0.98) |
| LDpred2-grid | P=0.01; $H^2$=0.05; S=FALSE | 0.35 (0.31, 0.40) | 0.98 (0.95, 1.00) |
| LDpred2-grid | P=0.1 $H^2$=0.05; S=FALSE | 0.32 (0.27, 0.37) | 0.98 (0.92, 1.00) |
| LDpred2-grid | P=1; $H^2$=0.05; S=FALSE | 0.31 (0.27, 0.36) | 0.99 (0.92, 1.00) |
| LDpred2-grid | P=0.001; $H^2$=0.1; S=FALSE | 0.11 (0.07, 0.16) | 0.96 (0.68, 0.98) |
| LDpred2-grid | P=0.01; $H^2$=0.1; S=FALSE | 0.33 (0.28, 0.37) | 1.00 (0.96, 1.00) |
| LDpred2-grid | P=0.1 $H^2$=0.1; S=FALSE | 0.34 (0.29, 0.38) | 0.98 (0.92, 1.00) |
| LDpred2-grid | P=1; $H^2$=0.1; S=FALSE | 0.33 (0.28, 0.37) | 0.99 (0.94, 1.00) |
| LDpred2-grid | P=0.001; $H^2$=0.15; S=FALSE | 0.09 (0.042, 0.13) | 0.90 (0.61, 0.95) |
| LDpred2-grid | P=0.01; $H^2$=0.15; S=FALSE | 0.33 (0.28, 0.37) | 1.00 (0.93, 1.00) |
| LDpred2-grid | P=0.1; $H^2$=0.15; S=FALSE | 0.33 (0.29, 0.38) | 0.99 (0.93, 1.00) |
| LDpred2-grid | P=1 $H^2$=0.15; S=FALSE | 0.32 (0.28, 0.37) | 0.99 (0.90, 1.00) |
| LDpred2-grid | P=0.01; $H^2$=0.20; S=FALSE | 0.28 (0.23, 0.32) | 0.99 (0.93, 1.00) |
| LDpred2-grid | P=0.1; $H^2$=0.20; S=FALSE | 0.33 (0.29, 0.38) | 0.98 (0.92, 1.00) |
| LDpred2-grid | P=1 $H^2$=0.20; S=FALSE | 0.33 (0.28, 0.37) | 0.98 (0.92, 1.00) |
| LDpred2-grid | P=0.001; $H^2$=0.05; S=TRUE | 0.14 (0.094, 0.18) | 0.93 (0.73, 0.98) |
| LDpred2-grid | P=0.01; $H^2$=0.05; S=TRUE | 0.36 (0.32, 0.41) | 1.00 (0.96, 1.00) |
| LDpred2-grid | P=0.1 $H^2$=0.05; S=TRUE | 0.33 (0.29, 0.38) | 0.98 (0.92, 1.00) |
| LDpred2-grid | P=1; $H^2$=0.05; S=TRUE | 0.33 (0.28, 0.37) | 0.99 (0.93, 1.00) |
| LDpred2-grid | P=0.001; $H^2$=0.10; S=TRUE | 0.10 (0.057, 0.15) | 0.94 (0.70, 0.98) |
| LDpred2-grid | P=0.01; $H^2$=0.10; S=TRUE | 0.34 (0.29, 0.38) | 1.00 (0.92, 1.00) |
| LDpred2-grid | P=0.1 $H^2$=0.10; S=TRUE | 0.35 (0.3, 0.39) | 0.99 (0.93, 1.00) |
| LDpred2-grid | P=1; $H^2$=0.10; S=TRUE | 0.32 (0.27, 0.36) | 0.98 (0.92, 1.00) |
| LDpred2-grid | P=0.001; $H^2$=0.15; S=TRUE | 0.1 (0.058, 0.14) | 0.94 (0.58, 0.96) |
| LDpred2-grid | P=0.01; $H^2$=0.15; S=TRUE | 0.31 (0.27, 0.36) | 0.99 (0.93, 1.00) |
| LDpred2-grid | P=0.1 $H^2$=0.15; S=TRUE | 0.35 (0.30, 0.39) | 0.99 (0.93, 1.00) |
| LDpred2-grid | P=1; $H^2$=0.15; S=TRUE | 0.33 (0.28, 0.37) | 0.99 (0.94, 1.00) |
| LDpred2-grid | P=0.01; $H^2$=0.20; S=TRUE | 0.28 (0.24, 0.32) | 0.99 (0.90, 1.00) |
| LDpred2-grid | P=0.1 $H^2$=0.20; S=TRUE | 0.34 (0.30, 0.38) | 0.99 (0.93, 1.00) |
| LDpred2-grid | P=1; $H^2$=0.20; S=TRUE | 0.31 (0.27, 0.35) | 0.98 (0.91, 1.00) |

P=P threshold for LDpred2-grid model; $H^2$=heritability; S=sparsity

**Supplementary Figure. P+T model selection in the *tuning dataset*.**

vPGS performance quantified using the Diff metric across a range of p-value thresholds, using vGWAS summary statistics from SCAMPI, conditional quantile regression (CQR), or Levene's test (LT).

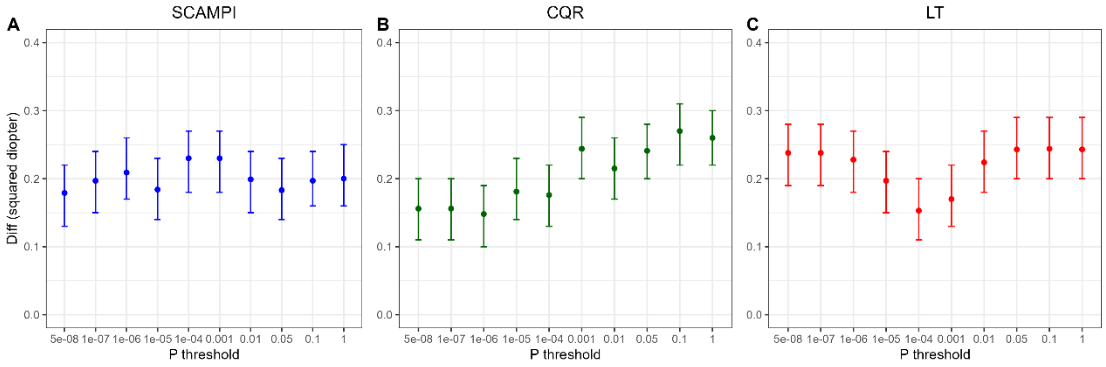
